# Physical health complaints among healthcare workers engaged in the care of critically ill COVID-19 patients: A single tertiary-care center prospective study from Japan

**DOI:** 10.1101/2020.10.09.20210393

**Authors:** Hiroki Namikawa, Yoshihiro Tochino, Akiko Okada, Keiko Ota, Yasuyo Okada, Kazuya Fujioka, Koichi Yamada, Tetsuya Watanabe, Yu Nakagama, Yasutoshi Kido, Yasuhiko Takemoto, Yasumitsu Mizobata, Hiroshi Kakeya, Yumiko Kuwatsuru, Toshihiko Shibata, Taichi Shuto

## Abstract

**Background:** Healthcare workers (HCWs) who care for patients with the novel coronavirus infectious disease (COVID-19) are at an increased risk and fear contracting the infection themselves. HCWs are chronically exposed to very intense stress, both and physically and mentally. Hospitals must reduce both the physical and mental burden of HCWs on the front lines and ensure their safety. No prospective study has focused on the physical health complaints among HCWs engaged in the care of critically ill COVID-19 patients. This study aimed to investigate the occupational risk among HCWs of experiencing physical symptoms during the current COVID-19 pandemic.

**Methods:** A twice-weekly questionnaire targeting HCWs who care for COVID-19 patients was performed at Osaka City University Hospital from April 30 to May 31, 2020 using a shareable Research Electronic Data Capture tool. The demographic characteristics of the participants, frequency of exposure to at-risk care, and physical complaints were evaluated.

**Results:** A total of 35 doctors, 88 nurses, and 35 technicians were engaged in the care of these critically ill COVID-19 patients. 76 HCWs participated in this study, of whom 24 (31.6%) were doctors, 43 (56.6%) were nurses, and 9 (11.8%) were technicians. The frequency of experiencing any physical symptom was 25.0% among HCWs. Exposure to at-risk care was significantly higher among nurses than among doctors (p < 0.001); likewise, the frequency of experiencing physical symptoms was higher among nurses than among doctors (p < 0.01). The multivariate analysis revealed that nurses (odds ratio 8.29; p = 0.01) might be independently at risk of experiencing physical symptoms.

**Conclusions:** Our results indicate that occupational health care at hospitals must be allocated to HCWs who are highly exposed to at-risk care, particularly nurses engaged in the care of COVID-19 patients.

## Introduction

The novel coronavirus disease 2019 (COVID-19) was first identified in Wuhan, Hubei Province, China, in late December 2019 [1]. It has since spread rapidly throughout the world, causing a global pandemic [2]. Consequently, this infectious disease is currently attracting the most attention in clinical practice. Healthcare workers (HCWs) with COVID-19 patients work very hard every day and are always at a high risk of infection, particularly when caring for critically ill patients with pneumonia caused by COIVD-19. Some studies have reported cases of HCWs infected with COVID-19 [3,4,5]. HCWs are chronically exposed to very intense stress, both and physically and mentally. Such an allostatic load may lead to disease over time [6]. Therefore, it is essential that hospitals reduce both the physical and mental burden of HCWs on the front lines and protect their safety.

A few clinical studies have reported on the management of mental health among HCWs during the COVID-19 pandemic [7,8,9]. However, to the best of our knowledge, no prospective studies have focused on the physical health care of medical workers caring for critically ill patients with COVID-19 pneumonia. The aim of this study was to investigate the association between occupation and the manifestation of physical symptoms among HCWs at a tertiary hospital in Japan during the current COVID-19 pandemic.

## Materials and Methods

### Study setting, population, and procedure

This prospective cohort study was performed from April 30 to May 31, 2020 using a shareable Research Electronic Data Capture (REDCap) tool [10]. During this study period, HCWs were actively involved in the care of critically ill patients with COVID-19 pneumonia who were admitted to Osaka City University. We defined patients who were admitted to the intensive care unit (ICU) and required mechanical ventilation as critically ill patients [11]. The study participants included doctors, nurses, and technicians (radiological and biomedical equipment technicians). Questionnaires were delivered to the study participants twice a week via REDCap. When the study participant did not perform patient care for 14 days in a row, we stopped delivering the questionnaire to them. Finally, we targeted the people who completed these questionnaires. The present study flowchart diagram is shown in Figure 1. This study was approved by the Ethics Committee of Osaka City University (approval number 2020-024). The purpose of this questionnaire was to manage the health of HCWs, which was different from the research purposes. Therefore, the need for written informed consent was waived owing to the clinical research using opt-out. This has been approved by the Ethics Committee of Osaka City University.

**Figure 1:**
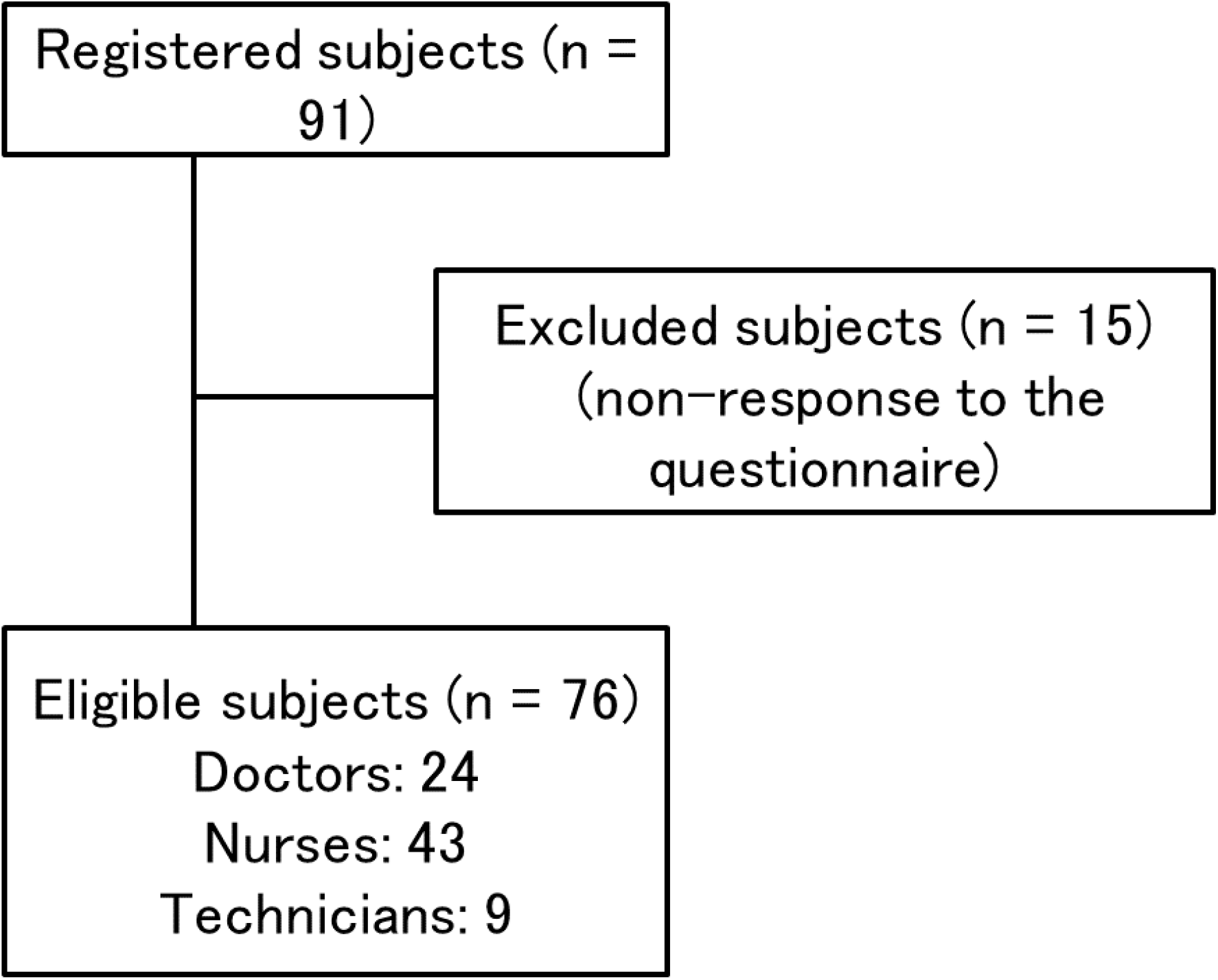
The present study flowchart diagram. 91 HCWs were enrolled in REDCap. Seventy-six HCWs were eligible for this study. Fifteen HCWs did not respond to the questionnaire. Of the 76 HCWs, 24 were doctors, 43 were nurses, and 9 were technicians.

### Screening questionnaire items

The questionnaire was developed for this study. The questionnaire assessed three main components: demographic characteristics, exposure, and physical complaints of postexposure (Supplementary Fig. S1). Demographic characteristics included age, gender, and occupation (doctor, nurse, or technician). Exposure included consecutive period and frequency (days a week, 1–7). Physical complaints consisted of attention-requiring and observation-requiring symptoms. Attention-requiring symptoms included fever, intense fatigue, dyspnea or shortness of breath, and dysosmia or dysgeusia (yes or no). Observation-requiring symptoms included sore throat, myalgia or arthralgia, headache, cough or sputum production, diarrhea or stomachache, nasal discharge or sneeze or nasal obstruction, and congestion or ophthalmalgia or low vision (yes or no). We also confirmed whether the participants requested an internal medical examination.

### Serological and molecular analysis

We performed antibody testing and real-time polymerase chain reaction (PCR) to identify severe acute respiratory syndrome coronavirus 2 (SARS-CoV-2) infection. We subjected HCWs engaged in the care of critically ill COVID-19 patients to antibody testing and those who were strongly suspected of having COVID-19 to PCR testing. We performed antibody testing two to four weeks after the HCWs were engaged. We used the SARS-CoV-2 IgM/IgG Quantum Dot immunoassay (Mokobio Biotechnology, MD, United States of America) for antibody testing according to the manufacturer’s instructions. SARS-CoV-2 real-time PCR was performed on the BD MAX ™ platform.

### Study outcomes

We evaluated the prevalence of physical symptoms of postexposure reported by the HCWs. Additionally, we investigated the association between occupation and the manifestation of physical symptoms among the HCWs.

### Statistical analyses

Participant characteristics, exposure, physical complaints, and laboratory data were compared between doctor, nurse, and technician groups. Fisher’s exact test and the Kruskal-Wallis test were used for the univariate comparison of categorical data. Additionally, the Bonferroni correction method was used for p value adjustment. To determine the independent predictors of the manifestation of physical symptoms, variables determined to be clinically important based on prior studies were considered for inclusion in the multivariate logistic regressions. All statistical analyses were performed with EZR [12], which is for R software. EZR is a modified version of the R commander that includes the statistical functions frequently used in biostatistics. A p value of <0.05 was considered statistically significant.

## Results

Eight (a cumulative total of nine: one patient was readmitted to the hospital after being discharged) critically ill patients with COVID-19 pneumonia were admitted to Osaka City University. All patients were male, with an average age of 63 years (38–81 years) and an average length of stay of 20 days (6–53 days). Two patients died while in the hospital. A total of 35 doctors, 88 nurses, and 35 technicians were engaged in the care of these critically ill COVID-19 patients. During our study period, 91 HCWs were enrolled in REDCap. Seventy-six HCWs were eligible for this study. Fifteen HCWs did not respond to the questionnaire. The baseline characteristics of the participants who were eligible for this study are summarized in Table 1. The 76 HCWs included 34 men and 43 women with a mean age of 36.2 years. Of the 76 HCWs, 24 (31.6%) were doctors, 43 (56.6%) were nurses, and 9 (11.8%) were technicians (radiological: 7, biomedical equipment: 2). Sixty-two (81.6%) HCWs were engaged for longer than two weeks. The participants reported a mean of 3.6 maximum exposure days per week. Nineteen HCWs (25.0%) had physical complaints, one (1.3%) had fever, four (5.3%) had intense fatigue, ten (13.2%) had headaches, and seven (9.2%) had nasal symptoms. A comparison of the baseline characteristics and laboratory findings between occupations is summarized in Table 2. There were significant differences in age (p < 0.01), exposure period (p < 0.001), physical complaints (p < 0.01), and headache (p < 0.01) among the three groups. Additionally, the nurses were noticeably younger than the doctors (p = 0.03), the nurses had a longer exposure period than the doctors (p < 0.001), and the frequency of physical symptoms was obviously higher in the nurses than in the doctors (p < 0.01). Conversely, no HCWs were suspected of being infected according to the laboratory findings. The multivariate analysis (explanatory variables: age, nurse, and exposure frequency) revealed that being a nurse (odds ratio 8.29; p = 0.01) might be an independent predictor of the manifestation of physical symptoms (Table 3). None of the HCWs developed COVID-19.

**Table 1.** Baseline characteristics of the study participants

|  |  |  |
| --- | --- | --- |
| 5 |  |  |
| 6 | Variables |  |
| 7 | Age (years)<sup>a</sup> | 36.2 ± 9.5 |
| 8 | Sex (male/female) | 34/42 |
| 9 | Occupation |  |
| 0 | Doctor | 24 (31.6%) |
| 1 | Nurse | 43 (56.6%) |
| 2 | Technician | 9 (11.8%) |
| 3 | Exposure |  |
| 4 | Consecutive period |  |
| 5 | ≤ 2 weeks | 14 (18.4%) |
| 6 | > 2 weeks | 62 (81.6%) |
| 7 | Frequency (days a week) |  |
| 8 | 1 | 6 (7.9%) |
| 9 | 2 | 15 (19.7%) |
| 0 | 3 | 19 (25.0%) |
| 1 | 4 | 15 (19.7%) |
| 2 | 5 | 13 (17.1%) |
| 3 | 6 | 2 (2.6%) |
| 4 | 7 | 6 (7.9%) |
| 5 | Physical complaint | 19 (25.0%) |
| 6 | Attention-requiring | 5 (6.6%) |
| 7 | Fever | 1 (1.3%) |
| 8 | Intense fatigue | 4 (5.3%) |
| 9 | Dyspnea or shortness of breath | 0 (0%) |
| 0 | Dysosmia or dysgeusia | 0 (0%) |
| 1 | Observation-requiring | 18 (23.7%) |
| 2 | Sore throat | 4 (5.3%) |
| 3 | Myalgia or arthralgia | 4 (5.3%) |
| 4 | Headache | 10 (13.2%) |
| 5 | Cough or sputum production | 5 (6.6%) |
| 6 | Diarrhea or stomachache | 6 (7.9%) |
| 7 | Nasal discharge or sneeze or nasal obstruction | 7 (9.2%) |
| 8 | Congestion or ophthalmalgia or low vision | 2 (2.6%) |
| 9 | Applicant for internal medical examination | 0 (0%) |

<sup>a</sup> Data are presented as means ± standard deviation

**Table 2.** Comparison of baseline characteristics and laboratory findings between occupations

| 4 | Variables | Doctor (n = 24) | Nurse (n = 43) | Technician (n = 9) | p value <sup>a</sup> |
| --- | --- | --- | --- | --- | --- |
| 5 | Age (years) <sup>b</sup> | 38.9 ± 9.3 | 33.3 ± 8.2 | 42.6 ± 10.3 | <0.01 <sup>c</sup> |
| 6 | Male | 22 (91.7%) | 3 (7.0%) | 9 (100%) | <0.001 <sup>d</sup> |
| 7 | Exposure |  |  |  |  |
| 8 | Consecutive period |  |  |  |  |
| 9 | > 2 weeks | 10 (41.7%) | 43 (100.0%) | 9 (100%) | <0.001 <sup>e</sup> |
| 0 | Frequency (days a week) <sup>b</sup> | 3.6 ± 2.0 | 3.8 ± 1.4 | 2.6 ± 1.2 | 0.07 |
| 1 | Physical complaint | 1 (4.2%) | 17 (39.5%) | 1 (11.1%) | <0.01 <sup>f</sup> |
| 2 | Attention-requiring | 1 (4.2%) | 4 (9.3%) | 0 (0%) | 0.82 |
| 3 | Fever | 0 (0%) | 1 (2.3%) | 0 (0%) | 1.00 |
| 4 | Intense fatigue | 1 (4.2%) | 3 (7.0%) | 0 (0%) | 1.00 |
| 5 | Dyspnea or shortness of breath | 0 (0%) | 0 (0%) | 0 (0%) | NA |
| 6 | Dysosmia or Dysgeusia | 0 (0%) | 0 (0%) | 0 (0%) | NA |
| 7 | Observation-requiring | 1 (4.2%) | 16 (37.2%) | 1 (11.1%) | <0.01 <sup>g</sup> |
| 8 | Sore throat | 0 (0%) | 4 (9.3%) | 0 (0%) | 0.42 |
| 9 | Myalgia or arthralgia | 1 (4.2%) | 3 (7.0%) | 0 (0%) | 1.00 |
| 0 | Headache | 0 (0%) | 10 (23.3%) | 0 (0%) | <0.01 <sup>h</sup> |
| 1 | Cough or sputum production | 0 (0%) | 5 (11.6%) | 0 (0%) | 0.23 |
| 2 | Diarrhea or stomachache | 1 (4.2%) | 5 (11.6%) | 0 (0%) | 0.47 |
| 3 | Nasal discharge or sneeze or nasal obstruction | 0 (0%) | 6 (14.0%) | 1 (11.1%) | 0.13 |
| 4 | Congestion or ophthalmalgia or low vision | 0 (0%) | 2 (2.6%) | 0 (0%) | 0.64 |
| 5 | Applicant for internal medical examination | 0 (0%) | 0 (0%) | 0 (0%) | NA |
| 6 | SARS-CoV-2 IgM positive<sup>i</sup> | 0 (0%) | 0 (0%) | 0 (0%) | NA |
| 7 | SARS-CoV-2 IgG positive<sup>i</sup> | 1 (5.0%) | 0 (0%) | 0 (0%) | NA |
| 8 | SARS-CoV-2 real-time PCR positive<sup>j</sup> | 0 (0%) | 0 (0%) | NA | NA |

9 <sup>a</sup> Continuous variable: Kruskal-Wallis test with the Bonferroni correction method
0 Categorical variable: Fisher's exact test with the Bonferroni correction method
<sup>b</sup> Data are presented as means ± standard deviation
<sup>c</sup> Doctor versus Nurse, p = 0.03, Doctor versus Technician, p = 0.97, Nurse versus Technician, p = 0.08
<sup>d</sup> Doctor versus Nurse, p < 0.001, Doctor versus Technician, p = 1.0, Nurse versus Technician, p < 0.001
<sup>e</sup> Doctor versus Nurse, p < 0.001, Doctor versus Technician, p = 0.01, Nurse versus Technician, p = 1.0
<sup>f</sup> Doctor versus Nurse, p < 0.01, Doctor versus Technician, p = 1.0, Nurse versus Technician, p = 0.42
<sup>g</sup> Doctor versus Nurse, p < 0.01, Doctor versus Technician, p = 1.0, Nurse versus Technician, p = 0.72
<sup>h</sup> Doctor versus Nurse, p = 0.03, Doctor versus Technician, p = 1.0, Nurse versus Technician, p = 0.53

**Table 3.** Multivariate analysis of predictors associated with the manifestation of physical symptoms

|  |  |  |  |
| --- | --- | --- | --- |
| 5 | Predictor | OR (95% CI) | p value |
| 6 | Age | 0.97 (0.90–1.04) | 0.40 |
| 7 | Nurse | 8.29 (1.67–41.2) | 0.01 |
| 8 | Exposure frequency | 1.23 (0.83–1.82) | 0.31 |

9 CI, confidence interval; OR, odds ratio

## Discussion

Our single-center, prospective cohort study of 76 HCWs caring for critically ill patients with COVID-19 pneumonia revealed that 25.0% of such HCWs developed physical symptoms. Second, our findings suggested that being a nurse may be an independent predictor of the manifestation of physical symptoms from working with COVID-19 patients.

The most common symptoms of non-severe COVID-19 are fever (43.0%–96.0%), cough (67.3%–71.0%), and fatigue (37.8%–39.0%); less common symptoms include headache (12%–13.4%), nasal congestion (5.1%), and diarrhea (3.5%–4.0%) [13,14]. Previous studies have indicated that SARS-CoV-2 might primarily target the lower respiratory tract [14,15]. Conversely, in our study, the most common symptoms in HCWs were headache (13.2%), nasal symptoms (9.2%), and gastrointestinal symptoms (7.9%); less common symptoms included fever (1.3%), intense fatigue (5.3%), and cough or sputum production (6.6%). We, as HCWs, must work under extreme pressure during the pandemic. Infectious disease outbreaks are known to have a strong psychological impact on HCWs. A recent study showed that HCWs facing excessive workloads and life-threatening conditions experienced psychological pressure and even worse mental illness during the SARS-CoV-2 outbreak [16]. Furthermore, psychological stress from the occurrence of infections can lead to both mental manifestations and various physical symptoms [17]. Recent studies have reported that there was a significant association between psychological outcomes and physical symptoms in HCWs during the current COVID-19 pandemic [17,18,19]. Chew et al. reported that common physical symptoms associated with the psychological impact of the COVID-19 outbreak among HCWs included headache (31.9%), throat pain (30.0%), joint/muscle pain (20.6%), cough (16.9%), coryza (14.0%), and sputum (11.3%) [17]. In fact, Ong et al. reported that the longer the duration of personal protective equipment (PPE) exposure among frontline HCWs during COVID-19, the more frequent they experienced headaches, which were known as PPE-associated headaches [20]. In light of the above findings, we believe that the various physical symptoms in our study were not caused by COVID-19 but rather by psychological stress and other factors including PPE exposure. One HCW tested positive for SARS-CoV-2 IgG in this study. Previous studies have reported that the incubation period for COVID-19 was less than 14 days, with most cases occurring within approximately five days of exposure [1,13]. Both IgM and IgG antibody levels against SARS-CoV-2 begin to increase from the second week after symptom onset [21]. Further, Xiao et al. reported that IgM levels almost disappeared by week seven, whereas IgG levels were persistently detectable beyond seven weeks [22]. In our study, this HCW did not complain of any physical symptoms while working with COVID-19 patients. Furthermore, this HCW underwent the SARS-CoV-2 IgM/IgG antibody test two weeks after beginning to work with COVID-19 patients and tested negative for SARS-CoV-2 IgM. Additionally, this HCW’s PCR testing result was negative. Based on the above findings, we considered the HCW to have had a prior infection with SARS-CoV-2 or a pseudo-positive test result, and consequently it was not considered a nosocomial infection.

In our study, the nurses tended to be younger and were more frequently exposed to COVID-19 patients than the other professions. Additionally, most of the nurses were women (93.0%). Liang et al. reported that younger (age ≤ 30) medical staff had higher self-rated depression scores than older medical staff during the COVID-19 outbreak [23]. Lai et al. showed that women had a higher risk of experiencing symptoms of distress among HCWs exposed to COVID-19 [24].

Further, the same study reported that nurses experienced more depression, anxiety, insomnia, and distress compared with doctors. Cai et al. also showed that nursing staff felt more nervous and anxious compared with other frontline medical staff during the COVID-19 outbreak [25]. A previous study reported that HCWs, particularly those working in emergency rooms, ICUs, and infectious isolation wards, are at a higher risk of developing adverse psychological outcomes [26]. HCWs are expected to be exposed to a significant degree of stress due to the COVID-19 pandemic and face various psychological symptoms. One important reason for the psychological distress experienced by HCWs exposed to patients with COVID-19 is the long-term workload [27]. Nurses, in particular, are likely to have many opportunities of intense contact with COVID-19 patients compared with other professions due to procedures involving extensive physical contact such as repositioning and suctioning. Therefore, we believe nurses are more likely to have physical symptoms related to the psychological stress. These findings indicate that being a nurse is an independent predictor of the manifestation of physical symptoms.

The present study has several limitations. First, the study population was relatively small, and the study was conducted in only one tertiary hospital. Therefore, there was selection bias. Future studies including more participants working with COVID-19 patients in both community and tertiary hospital settings are required to address this limitation. Second, physical assessment was based on a self-reported online survey using a REDCap tool. Physical examination is recommended in future studies to conduct a more detailed assessment of the physical problems. Third, we examined 11 physical complaints and performed serological and molecular biological analyses, but we did not examine psychological problems in HCWs caring for COVID-19 patients. Therefore, we are unable to conclude whether the manifestation of these physical symptoms resulted from psychological distress. Psychological problems should be analyzed to assess the association between these physical symptoms and psychological outcomes.

In summary, the present study demonstrated that the frequency of physical symptoms in HCWs caring for critically ill patients with COVID-19 pneumonia was 25.0%. Being a nurse may be an independent predictor of the manifestation of physical symptoms from working with COVID-19 patients. Therefore, we must manage the health care of both the physical and mental symptoms of HCWs, particularly nurses, who work with COVID-19 patients. We believe that this study is the first step toward establishing a physical health management strategy for HCWs with COVID-19 patients.

## Data Availability

None

## Authors’ contributions

NH, TYO, and STO designed this study. OA and OK set up the survey system. FK, NY, and KYA conducted the serological and molecular analysis. OY, YK, WT, MY, KH, KYU, and STO conducted the infection management in our hospital. NH, TYO, YK, TYA, KH, STO, and STA conducted the clinical interpretation. NH and TYO drafted the manuscript and critically revised it. All authors contributed to the final version of the manuscript and approved its submission.

## Acknowledgments

We thank Hisako Yoshida (Department of Medical Statistics, Osaka City University, Graduate School of Medicine) for her technical support and advice in medical statistics.

## Additional files

**Supplementary Figure 1:**
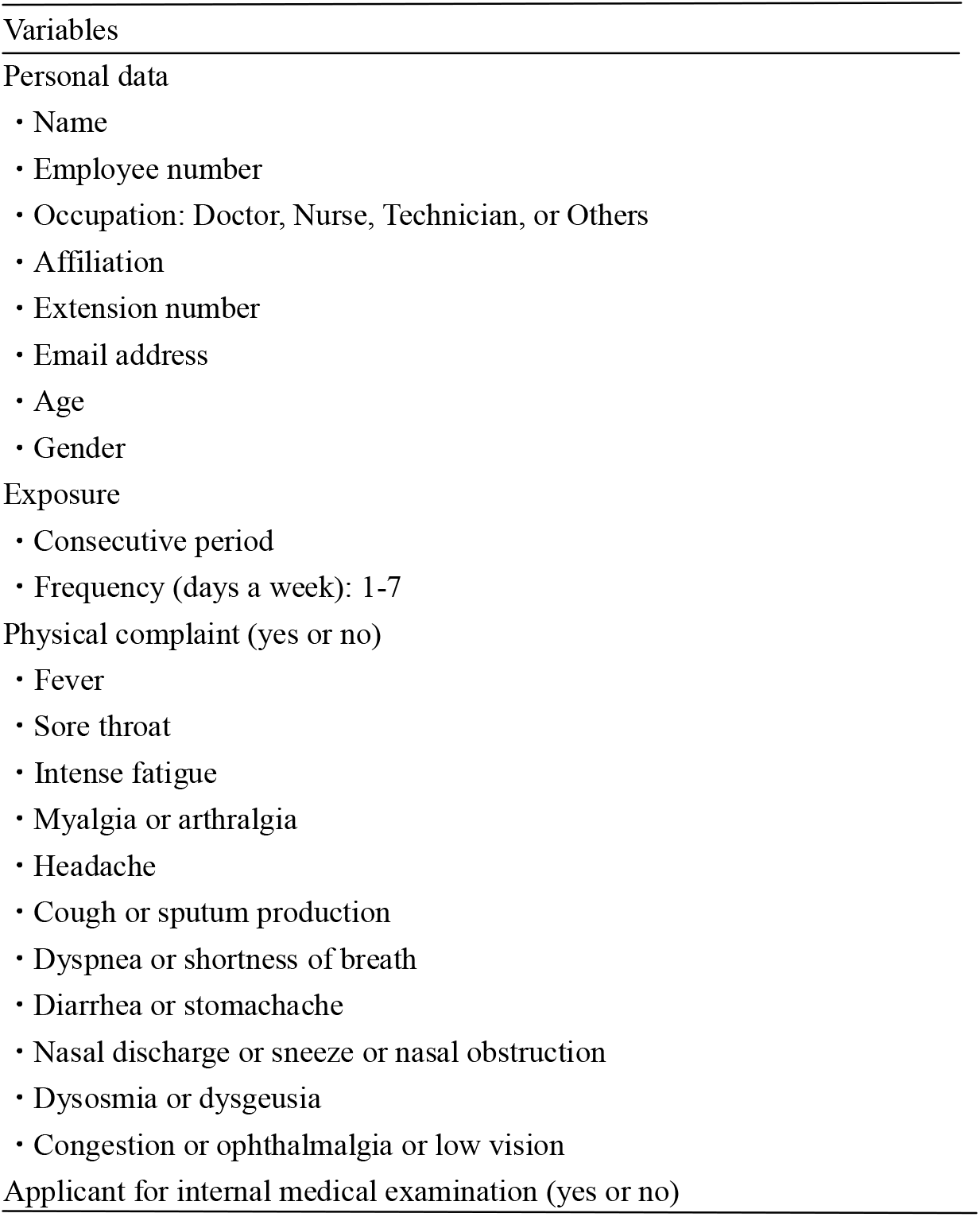
Questionnaire details. The questionnaire assessed three main components: demographic characteristics, exposure, and physical complaints of postexposure. We also confirmed whether the participants requested an internal medical examination.

